# Optimal timing of one-shot interventions for epidemic control

**DOI:** 10.1101/2020.03.02.20030007

**Authors:** Francesco Di Lauro, István Z. Kiss, Joel C. Miller

## Abstract

The interventions and outcomes in the ongoing SARS-CoV-2 pandemic are highly varied. The disease and the interventions both impose costs and harm on society. Some interventions with particularly high costs may only be implemented briefly. The design of optimal policy requires consideration of many intervention scenarios. In this paper we investigate the optimal timing of interventions that are not sustainable for a long period. Specifically, we look at at the impact of a single short-term non-repeated intervention (a “one-shot intervention”) on an epidemic and consider the impact of the intervention’s timing. To minimize the total number infected, the intervention should start close to the peak so that there is minimal rebound once the intervention is stopped. To minimise the peak prevalence, it should start earlier, leading to initial reduction and then having a rebound to the same prevalence as the pre-intervention peak rather than one very large peak. To delay infections as much as possible (as might be appropriate if we expect improved interventions or treatments to be developed), earlier interventions have clear benefit. In populations with distinct subgroups, synchronized interventions are less effective than targeting the interventions in each subcommunity separately.

**Author Summary:** Some interventions which help control a spreading epidemic have significant adverse effects on the population, and cannot be maintained long-term. The optimal timing of such an intervention will depend on the ultimate goal.

- Interventions to delay the epidemic while new treatments or interventions are developed are best implemented as soon as possible.
- Interventions to minimize the peak prevalence are best implemented partway through the growth phase allowing immunity to build up so that the eventual rebound is not larger than the initial peak.
- Interventions to minimize the total number of infections are best implemented late in the growth phase to minimize the amount of rebound.

For a population with subcommunities which would have asynchronous outbreaks, similar results hold. Additionally, we find that it is best to target the intervention asynchronously to each subcommunity rather than synchronously across the population.

## 1 Introduction

The Influenza pandemic of 1918 was one of the deadliest epidemics of infectious disease the world has ever seen. In response, many cities introduced widespread interventions intended to reduce the spread. There is evidence [6] that some cities which implemented these interventions later had fewer deaths. This seemingly counter-intuitive observation suggests that they were more successful by being slow to respond.

When the 2009 influenza pandemic first arrived outside of Mexico, many schools shut after the first observed infection. Once these schools reopened, and received a new introduction, the remaining susceptible population was almost as large as at the outset, so the resulting epidemic was likely to be nearly as large as the original epidemics would have been. The closure provided increased time to prepare a response and learn more about the disease, but the overall epidemic was very similar to what would have happened without the closure. In contrast, evidence suggests that summer holidays altered the final outcome of the pandemic (at least in the UK), significantly reducing the total number of infections by splitting the epidemic into two smaller peaks [13].

The phenomenon can be explained by noting that epidemics rely on two things to spread: infected individuals and a supply of susceptible individuals. If the intervention is too early, the number infected may fall, but there will be enough susceptibles available that it can re-establish and grow again. When it returns to the original size, the remaining susceptible population will be effectively the same size as it was the first time. Thus, the intervention primarily delays the spread; the resulting epidemic is comparable to what would have been seen before. However, if the intervention occurs once the susceptible population has been noticeably depleted, then the number of infections falls and when the intervention is relaxed, the depleted susceptible population makes the rebound smaller or even nonexistent.

To make this explanation more robust, we note that is well-known that after an unmitigated epidemic, the total number of infections exceeds the number of infections required to achieve the “herd immunity threshold” (the level of immunity required to reduce the effective reproduction number below one) [8]. We refer to this extra level of infection as the “overshoot”. It is a consequence of the fact that when the effective reproduction number (in absence of intervention) finally falls to 1, the population reaches the “herd immunity threshold” and incidence no longer increases. However, because the epidemic is at its peak, this is the time at which those who have escaped infection so far face the highest force of infection. As the number infected falls, significant transmission still happens and the epidemic overshoots the herd immunity threshold. In the absence of further intervention, the size of the overshoot is determined by the number infected when the herd immunity threshold is reached.

When we think about this in terms of a temporary intervention, the option to minimize the total number of infections becomes clearer. A short intervention that ends with the effective reproduction above one would see a rebound and would see a larger overshoot than a slightly later intervention that ends with the effective reproduction number equal to 1.

This underlies the explanation of [6] for why temporary interventions are generally more effective if introduced later in the epidemic (but not too late). Similar, more detailed theoretical results have been found by [19, 1]. Most studies of these effects are focused on a single population, and they do not carefully consider the tradeoffs between competing goals of delaying infections, reducing the peak prevalence, or reducing the total size.

In the ongoing COVID-19 epidemic, China introduced drastic control measures very early. These significantly reduced transmission, apparently reducing the effective reproduction number (the number of new infections per infected individual) below one [23, 39], although it took a very long time for new cases to stop. Despite quite significant interventions in Italy in force for a long period of time, the rate of new cases was slow to fall [9].

Many other places have turned to aggressive control of infection in an attempt to keep transmission suppressed [20, 24, 18, 33, 37, 3, 22].

In places which have have nearly eliminated the disease, the threat of re-emergence requires constant vigilance. In places which have failed to contain transmission, the pervasive interventions that would be required to get transmission low would impose significant costs through the entire population, and such extensive interventions are unlikely to be maintained long term. Thus policy-makers face challenges about whether or when to implement such restrictive interventions.

Motivated by ongoing decisions facing policy makers for the COVID-19 pandemic, we develop mathematical models which allow us to explore how to time short-term interventions in response to an emerging epidemic. We will refer to these temporary interventions as “one-shot” interventions, meaning that the intervention cannot be maintained indefinitely or repeated. We are particularly interested in how the timing might affect the total fraction infected and the peak prevalence, but we are also interested in the resulting delay of infections.

We must exercise care in determining that a given intervention cannot be sustained. In the initial phase of the pandemic there was significant uncertainty in the fatality rate. With this in mind [38], the tolerance of the population for drastic interventions could be significant. What might appear to be an unsustainable intervention given one set of assumptions about severity may in fact be sustainable under another set of assumptions. We assume perfect information and focus on choosing the time at which a given strategy will be more effective. A separate, but related line of research focuses on whether (and how long) we should hold an intervention in reserve while we learn more about the disease: sometimes the greatest expected benefit comes from learning more before choosing the intervention [5, 26, 32]. For an epidemic that grows quickly (like the early stage of the SARS-CoV-2 pandemic), there is effectively no time to learn about the disease before a decision is needed, and so these strategies would not be relevant until strong enough interventions are in place to suppress transmission.

We model an infection spreading in an initially fully susceptible population. We will model the spread within a single well-mixed population and a population made up of several weakly-coupled subcommunities (a *metapopulation*). We will investigate the impact of intervention on the attack rate (the final fraction infected), the peak prevalence, and the timing of infections, and in the metapopulation model we will additionally consider whether it is better to have a synchronized intervention or to have the intervention timed separately for each subcommunity. The important question of whether disease can be eliminated locally is beyond our scope.

Our goal is not to provide predictions for a specific population, but rather to demonstrate the generic impact of delaying a one-shot intervention, to show its robustness, and to provide intuition and some guiding principles which will apply to more complex scenarios.

Our results have important implications for the ongoing COVID-19 epidemic. If an intervention cannot be sustained for an extended period of time but new interventions or treatments are being developed, it is likely to be best to perform the intervention sooner to delay potential infections until other methods are available to treat or further delay infection (e.g., masks distributed, contact tracing implemented, healthcare capacity increased, therapeutic treatments identified, or even vaccine produced). However, if no other intervention or treatment improvement is likely to emerge, then it is best if the intervention is “held in reserve” until depletion of susceptibles has reduced the effective reproductive number enough that the intervention will have maximal impact on the total number of infections (by preventing the overshoot) or the peak prevalence.

We completed and released this research early in the initial stages of the pandemic (at the end of February 2020) [11], but did not immediately pursue it further due to other pressing questions. In the interim, a number of other papers have emerged studying related questions, including [17, 14, 10, 15, 30]. It is clear from much of this work that nonpharmaceutical interventions are an important part of epidemic control. In particular, the timing of an intervention, be it in a single population or over different communities, has a major impact on its effectiveness and overall outcome.

Our results provide insights into ongoing discussions of “circuit-breaker” interventions: in particular, such an intervention particularly valuable because it can delay infections while other interventions are brought into place, and it can keep the infection count low enough that interventions that cannot scale well can remain effective. However, if there is no significant effort to increase other interventions, then a repeated sequence of such “circuit-breakers” may be needed or the circuit-breakers should be delayed.

In this paper, we first introduce the mathematical models we use to explore the impact of a one-shot intervention against an infectious disease in a single well-mixed population and in a metapopulation made up of several distinct subcommunities. Then we discuss results from those mathematical models. Finally we discuss the implications of these results. In the Appendix we develop some mathematical theory explaining the mechanism underlying the effect in more detail.

## 2 Methods

In this section we introduce mathematical models for an “SIR” (Susceptible–Infected–Recovered) epidemic in a single well-mixed population and in a metapopulation made up of several sub-communities. We assume that the intervention is initiated at a specific time *t*^*^ (typically once the cumulative number of infections *I* + *R* reaches some threshold), and that the intervention lasts for a fixed duration *D*. It reduces the transmission rate by a “strength factor” *c*. We explore the impact of the threshold, duration and strength of intervention. In the metapopulation model, we compare outcomes when the intervention is implemented in all populations at the same time or in each individual population separately. In both models we measure time in multiples of the typical infection duration.

We will measure the impact of interventions on three quantities of interest:

- the *attack rate* or *final size*: the total fraction infected *R*(∞),
- the *peak prevalence* or maximum value of *I*(*t*), and
- the *average time of infection*, 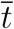, the average time (or date) at which individuals become infected. It is given by 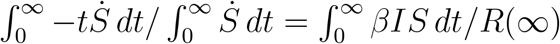.

In general the (often conflicting) goals of our intervention are to reduce *R*(∞), reduce *I*_max_, and increase 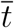.

The value of minimizing the attack rate is clear as it minimizes the number of infections. The value of minimizing the peak prevalence is highlighted by the struggles that many health systems have faced during the early phase of the COVID-19 pandemic. The importance of increasing the average date at which infections occur is somewhat less clear. However, early in an epidemic, medical knowledge about the disease, health care capacity, and testing/contact tracing capacities are likely to be limited. Important knowledge about the transmission mechanisms may be missing. In this early stage where knowledge is increasing, any intervention that delays the bulk of infections until later is likely to help increase both the quality of medical care provided and the effectiveness of interventions that may prevent those infections altogether in the future. This may be particularly important for interventions such as contact tracing which take time to put in place and lose effectiveness when there are many infections.

### 2.1 Well-mixed population

To study an intervention in a well-mixed population, we use the standard SIR model [2].

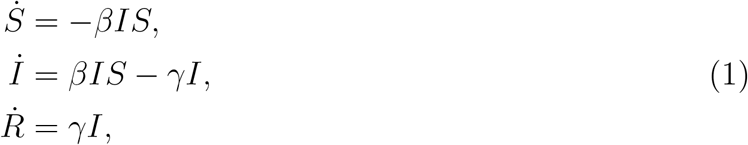

where *S, I*, and *R* denote the susceptible, infected and recovered fractions of the population with *S* + *I* + *R* = 1. There are a few important quantities to consider.

- The *basic reproduction number* ℛ_0_: The average number of infections an infected individual causes early in the epidemic in the absence of intervention and the absence of any depletion of susceptibles. This is ℛ_0_ = *β/γ*.
- The *effective reproduction number* ℛ_*e*_: As depletion of susceptibles occurs or interventions are put into place, the number of infections an infected individual causes is reduced. When ℛ_*e*_ < 1, the number of infections declines.

By measuring time in multiples of the typical infection duration, we impose that *γ* = 1, and so *β* = ℛ_0_.

If ℛ_0_ > 1 the typical behavior of an epidemic without an intervention is that at *t* = 0 we have *S* ≈ 1, *I* is very small and *R* = 0. As time increases, *I* and *R* grow and *S* decreases. The reduction in *S* reduces the effective reproduction number: ℛ_*e*_ = ℛ_0_*S*. Once *S* < 1*/*ℛ_0_, *I* begins to fall because recoveries outweigh new infections: *I* → 0. Some fraction remains uninfected: *S*(∞) > 0 and *R*(∞) = 1 − *S*(∞) [2, 27, 29]. See Figure 1 for typical profiles of *S, I*, and *R* in time.

**Figure 1:**
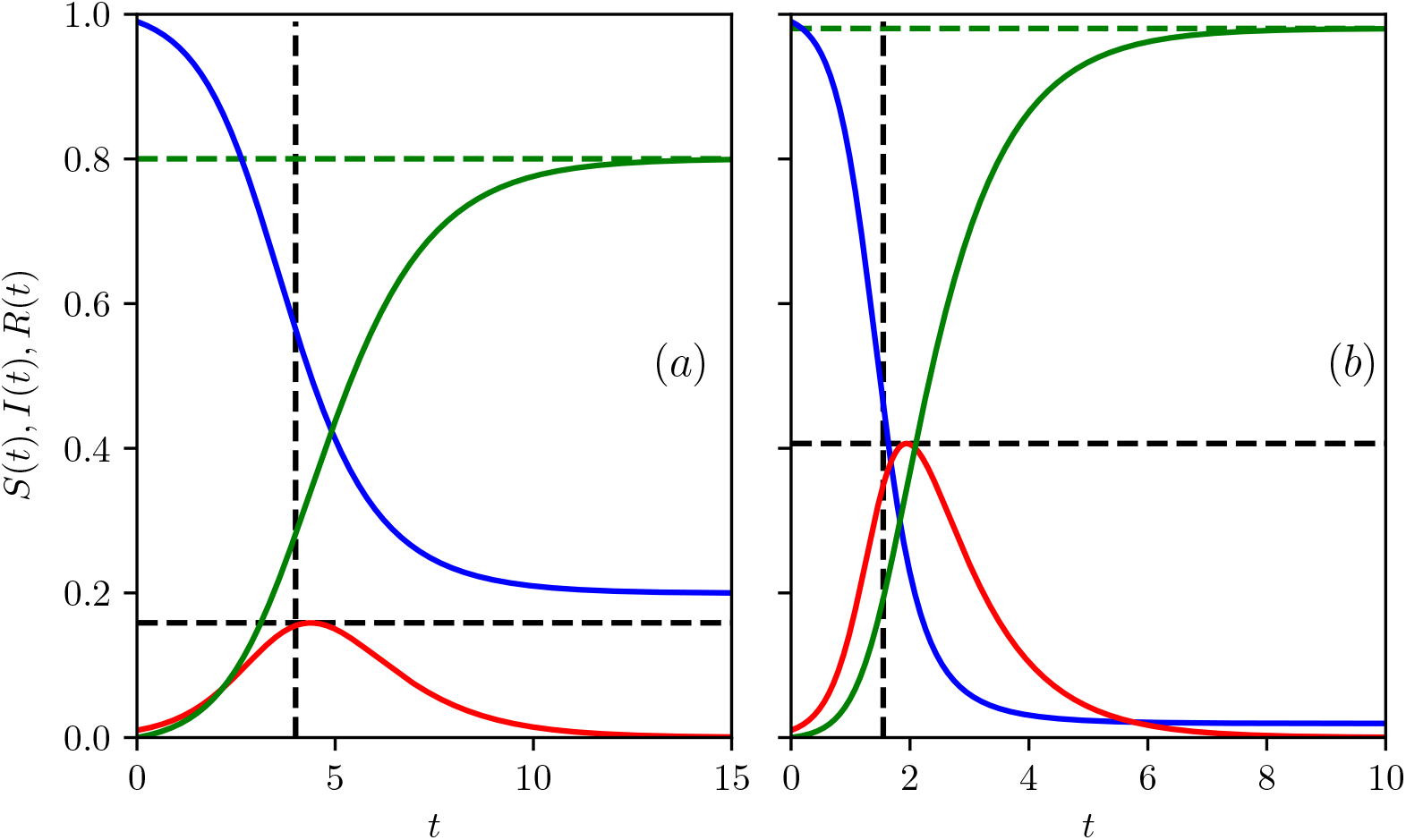
The time-evolution of *S, I* and *R* for epidemics with no control. (a) ℛ_0_ = *β* = 2 and (b) ℛ_0_ = *β* = 4 with *γ* = 1 in both. Horizontal and vertical dashed black lines indicate the peak prevalence *I*_max_ and average time of infection 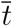 respectively, while green dashed horizontal lines show the attack rate *R*(∞) found by numerically solving 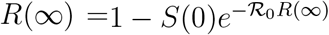.

We assume that at some time *t* = *t*^*^, an intervention that reduces the transmission rate is introduced for a duration *D*. The intervention reduces *β* by some factor *c*. So from time *t* = *t*^*^ to time *t* = *t*^*^ + *D* the transmission rate *β* = ℛ_0_ is replaced by *β* = (1 − *c*)ℛ_0_. During the intervention, the effective reproduction number is ℛ_*e*_ = *S*(1 − *c*)ℛ_0_. After time *t* = *t*^*^ + *D* the transmission rate returns to *β* = ℛ_0_, and ℛ_*e*_ = *S*ℛ_0_.

We will typically assume that *t*^*^ is chosen based on the cumulative number of infections *I*(*t*) + *R*(*t*) crossing some threshold. We choose a monotonically increasing measure *I* + *R* because this lets us choose an arbitrary *t*^*^, which would not be possible if we focused on prevalence (*I*) or instantaneous rate of infection 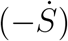.

### 2.2 Weakly-coupled Metapopulation model

We will also investigate the effectiveness of interventions in a metapopulation made up of distinct subcommunities that do not have synchronized epidemics. The most obvious reason for this setup would be geographically separated populations. However there could be stratification by age, religion, ethnicity or socio-economic status. We are particularly interested in whether it is better to time interventions to the dynamics within each subcommunity separately or for the intervention to be synchronized even through the respective epidemics are not.

It is well-known that if the subcommunities have strong enough coupling, the epidemics in all subcommunities are effectively synchronised [4, 12]. In this case there is little distinction between asynchronous interventions for each subcommunity or interventions synchronized across all subcommunities. Thus to compare the results from synchronized interventions with asynchronous interventions targeted to each subcommunity, we need to explore a population with weak coupling. We use a standard meta-population model [2], allowing most transmission to be within a subcommunity and some cross interactions between the sub-communities.

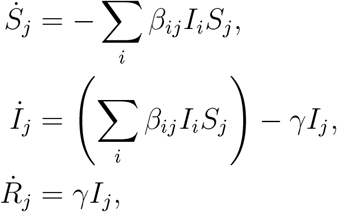

where 0 ≤ *S*_*i*_ ≤ 1, 0 ≤ *I*_*i*_ ≤ 1 and 0 ≤ *R*_*i*_ ≤ 1, with *S*_*i*_ + *I*_*i*_ + *R*_*i*_ = 1 for all *t*, represent the fraction of susceptible, infected and infectious and recovered individuals in subcommunity *i*, where *i* = 1, 2,…, *N*.

To simplify the presentation, all subcommunities are of equal size. The recovery rate *γ* is identical for all subcommunities. As before we measure time in multiples of the typical infectious period, so we set *γ* to 1. The cross-infection between subcommunities is modelled by *B* = (*β*_*ij*_)_*i,j*=1,2,…*N*_, where *β*_*ij*_ represents the rate at which infectious contacts are made from subcommunity *i* towards susceptible individuals in subcommunity *j*.

We implement a weak coupling by joining the population in a linear fashion: population *i* is only connected to population (*i* − 1) and (*i* + 1). The first and the last populations only connect to the second and the pen-ultimate population, respectively. The entries for the coupling/mixing matrix are generated as follows. On the main diagonal, the *β*_*ii*_ values are set to 2 + (Unif(0, 1) − 0.5) where Unif(0, 1) produces a random number chosen uniformly between 0 and 1. Off-diagonal entries are set to Unif(0, 1)(*β*^*^*/*10) (*β*^*^ = max_*i*=1,2, …,*N*_ *β*_*ii*_) and represent a scaled and randomised version of the largest entry on the main diagonal. This yields an ℛ_0_ above 2, comparable to current estimates for COVID-19 [23, 25].

We will use this model to explore whether it is better to implement an intervention in a synchronized fashion across all subcommunities or to implement it in each subcommunity. In particular, we will consider the following scenarios:

- track *I*_*i*_ + *R*_*i*_ in each subcommunity and as soon as *I*_*i*_ + *R*_*i*_ > *T* for some threshold *T*, a one-shot control is deployed in the corresponding subcommunity,
- track 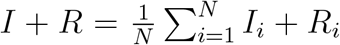 globally and as soon as *I* + *R* > *T*, a one-shot control across all subcommunities, and
- track each subcommunity and deploy the one-shot control is deployed across *all* sub-communities as soon as *I*_*i*_ + *R*_*i*_ > *T* for the first subcommunity.

One-shot control in a subcommunity is understood to mean reduction in the internal, incoming, and outgoing rates of infection with a factor of (1 − *c*), where 0 ≤ *c* ≤ 1 denotes the intervention strength (we assume that the strength is the same in each subcommunity, and if two communities are both acting, then the movement between them is scaled by (1 − *c*)^2^). This reduction lasts for a duration *D* and, as soon as the control is over, the transmission rates for that subcommunity are restored to the starting levels.

In our results, we will present the average outcome of simulations across 100 distinct populations whose mixing matrices are chosen stochastically based on the rules described above.

## 3 Results

We use our mathematical models to explore how the timing of a one-shot intervention can impact

- total attack rate,
- peak prevalence, and
- average time of infection.

These are expected to be good proxies of the total impact on the population or the burden on the health services.

We find that one-shot interventions that begin at the first sign of infection have the most impact on delaying the epidemic, but they have little impact on the attack rate or the peak prevalence. This is because only a few individuals are infected when the intervention is implemented so not many transmissions are blocked. When the restrictions are lifted, almost as many transmissions end up happening: the disease spreads in an almost fully susceptible population, and its trajectory is very nearly the same, just delayed. In contrast if the intervention is delayed until a non-negligible fraction of the population has been infected it will have more impact on the epidemic’s shape.

For the weakly-coupled metapopulation model, the subgroups are likely to have somewhat asynchronous epidemics. In this case it is better to implement the one-shot interventions based on a local threshold rather than a global threshold. If the coupling is stronger, the epidemics are closely synchronized and there is little difference between the strategies.

### 3.1 Well-mixed population

We can think of a strong, but temporary, intervention as dividing the overall epidemic into two phases. We allow an epidemic to spread until the intervention is started. The intervention resets *I* to a small value (that is, the intervention shifts the epidemic to a new trajectory with a similar *S*, but a smaller *I*). Depending on how long the epidemic was allowed to spread prior to the intervention, we have some new value of *S*(*t*^*^ +*D*). Then a new epidemic happens starting from the new initial state, spreading as if a fraction 1 − *S*(*t*^*^ + *D*) were vaccinated. The longer we allow the first phase epidemic to spread, the smaller the value of *S*(*t*^*^ + *D*), and so the smaller the second phase will be. An early intervention truncates the first phase, but a later intervention reduces the second phase.

For a well-mixed population we find that the timing of a one-shot intervention has an important impact on the total epidemic. If the intervention is put in place very early, then the impact is to simply delay the epidemic. Because *S*(*t*^*^ + *D*) ≈ *S*(0) the second phase is effectively like the first phase without an intervention, but delayed. The delay is somewhat larger than *D* because it takes some time for *I* to grow back to *I*(*t*^*^).

Figure 2 shows the impact of an intervention in a population with ℛ_0_ = 2.5 and an intervention of strength *c* = 4*/*5 (it prevents 4 of every 5 transmissions), and duration 2 (time units measured in multiples of the typical infection duration). The figure focuses on the impact of varying the threshold value of *I* + *R* at which the intervention is introduced.

**Figure 2:**
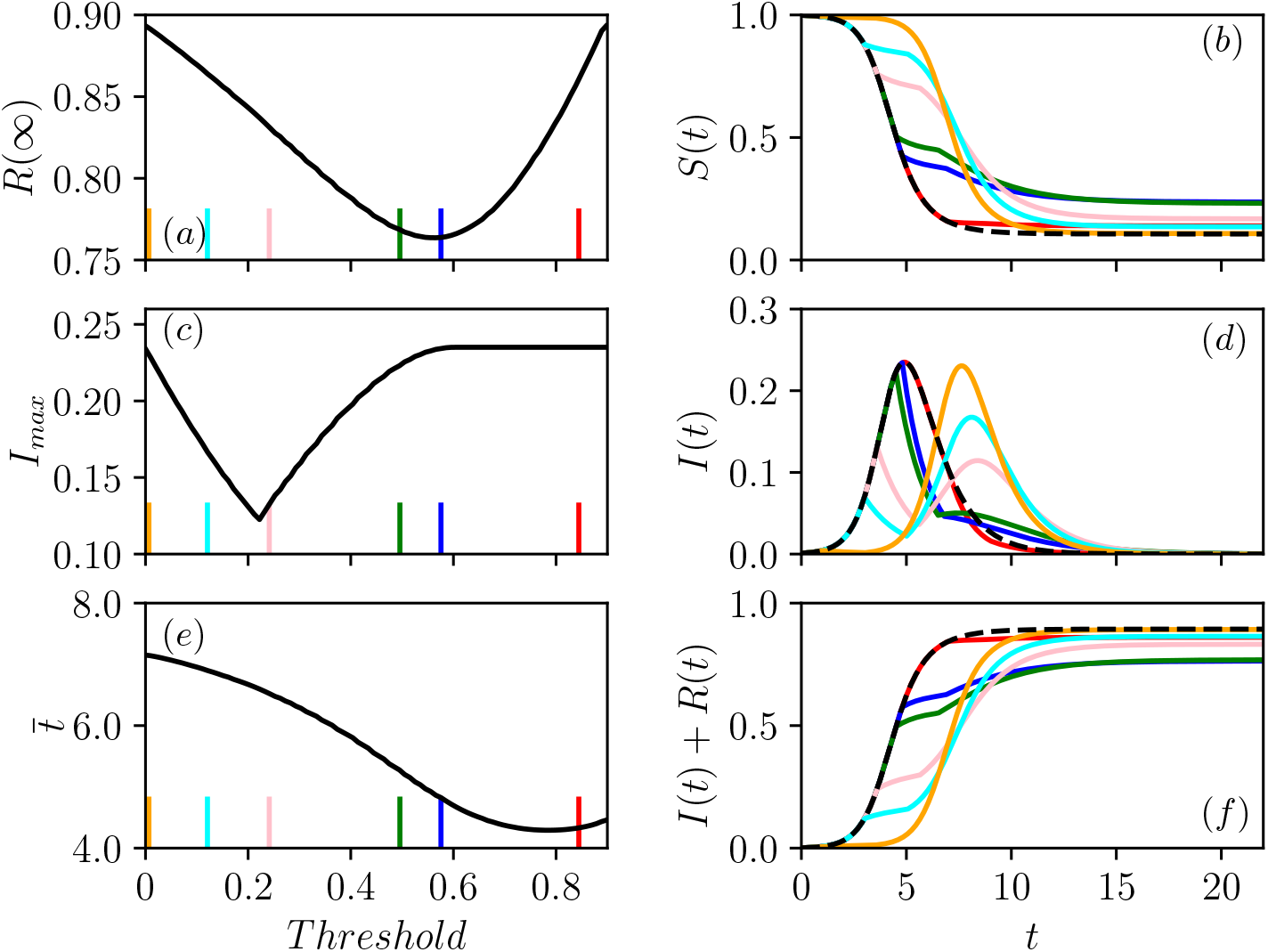
Illustration of the impact of one-shot intervention in a population with ℛ_0_ = 2.5. The intervention has *c* = 0.8 for a duration of *D* = 2 time units. This intervention is introduced at different times as determined by a range of *Threshold* values. The impact of the threshold (*I* +*R* > *Tr*) for implementing the intervention is shown for (a) the attack rate *R*(∞); (b) *S*(*t*); (c) peak prevalence *I*_max_; (d) *I*(*t*); (e) average time of infection 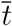; and (f) plots of *I*(*t*) + *R*(*t*). In (b,d,f), the no-control case is plotted as a dashed line. The vertical lines in (a,c,e) correspond to the threshold for cumulative infections *I* + *R* which yields the intervention leading to the corresponding color in (a,c,e).

Figure 3 shows how the optimal threshold changes as the parameters of the disease or intervention change.

**Figure 3:**
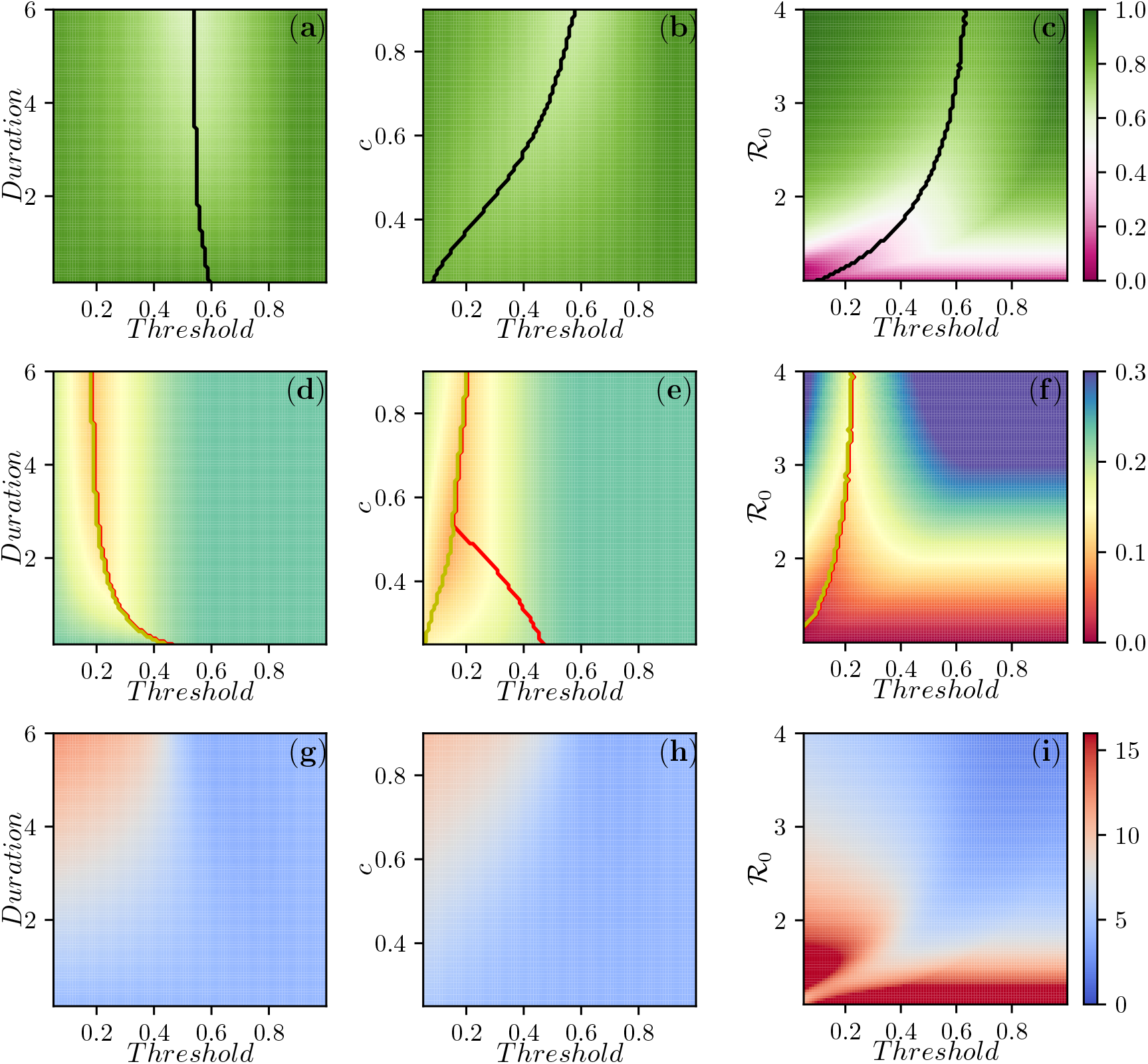
Contour plots for *R*(∞) (top), *I*_max_ (middle) and the mean time of infection 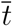 (bottom) as a function of parameters for the well-mixed population. We explore different threshold values of *I* + *R* for the intervention to start, from a minimum of 0.05 to a max of 0.9. The first column investigates the impact of duration from *D* = 0.1 to *D* = 6, holding *β* = 2.5 and *c* = 0.8. On the second column, intervention duration is *D* = 4 and *c* ranges from 0.2 to 0.9. Finally, on the third column, *c* = 0.8 and *D* = 4, and the values of *β* = ℛ_0_ vary from 1 to 4. In all cases *γ* = 1. In the first row, the black curve denotes the threshold for which ℛ_*e*_ = 1 when the intervention completes. In the three regions defined by the two lines in the panels of the second row, the peak prevalence is observed after the intervention has ended (from left to yellow curve), during intervention (area between the curves), or before intervention (from red curve to the end of the figure). Where the two curves align, the prevalence decays as soon as the intervention is implemented and then recovers to the pre-interention peak.

#### 3.1.1 Impact on attack rate

The impact on the attack rate (the total number infected) can be understood by a mental model of the intervention as a way to shift from the current epidemic trajectory to a new epidemic trajectory with a similar number of susceptibles, but fewer infected (this is made more rigorous in Appendix A.1).

If the intervention is introduced early on, it will have an immediate impact. However, when the intervention is lifted, the epidemic rebounds until the number of infections is the same as the original value. The number susceptible is relatively unchanged, and so a similar epidemic happens with an almost identical epidemic curve once it rebounds, except with a shift to later time. So an early intervention has little impact on the attack rate *R*(∞).

In Figure 2(a) we see that if the intervention is introduced later, there is clear improvement in *R*(∞), up to a threshold of *I* + *R* of 0.6, which is close to where the peak prevalence occurs in the epidemic without intervention. This is because when the epidemic peaks, ℛ_*e*_ = 1, and so if we immediately and dramatically reduce the number infected at this point the epidemic quickly dies out.

As new infections do happen during the intervention, this mental model is only an approximation. It can be made more precise by recognizing that to reduce the attack rate *R*(∞), the intervention is most effective if it is timed to directly block as many transmissions as possible. So we want to time the intervention to maximize the number of infected individuals present while it is in place (mathematically we want to maximize 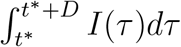 given *D, c*, and ℛ_0_).

Thus the ideal timing to reduce the total number of infections is not at the first hint of infection (when there are not enough infected individuals to cause many transmissions), but rather, a little before the peak, and if the intervention is perfect (*c* = 1), then at the peak. This suggests that the more effective an intervention is, the closer we should be to the peak before implementing it. It also suggests that for an intervention of a longer duration, we can implement it somewhat sooner, but not significantly sooner. For a more infectious disease, the need to begin the intervention near the peak implies that the threshold value of *I* + *R* will need to be larger (though the time *t*^*^ at which it is implemented is smaller).

These predictions are borne out by observations of the first column of Figure 3 which shows how the optimal threshold value of *I* + *R* for implementing the intervention changes as the strength *c*, the duration *D*, or the reproductive number ℛ_0_ change. The earliest interventions have the most impact on the average time of infection, while somewhat delayed interventions affect the peak prevalence the most, and later interventions (near the epidemic peak) affect the final attack rate.

#### 3.1.2 Impact on peak prevalence

As in the attack rate case, an early intervention primarily delays the epidemic curve. It does not significantly alter the shape. Thus the peak prevalence remains effectively the same unless the intervention is delayed until *S* is noticeably depleted.

If the susceptible population has been sufficiently depleted prior to the elimination of the intervention, then once the intervention is stopped, the epidemic rebound will be muted. Moving the intervention later makes the rebound smaller still. However, it means that the number of infections prior to the intervention is larger. There comes a threshold at which the phase before and the phase after the intervention have the same maximum. This is the time that minimizes the peak prevalence. Delaying the intervention past this value results in a larger pre-intervention peak, while doing it sooner results in a larger post-intervention peak.

Figure 2(c) shows the optimal threshold to reduce the peak prevalence occurs sooner than to reduce the attack rate. We can understand this intuitively, because for optimizing peak prevalence a moderate rebound is less of a concern than for optimizing the attack rate. For the purpose of reducing peak prevalence, Figure 2(d) shows that the optimal time to introduce the intervention is when the current prevalence matches the peak prevalence that would occur once the disease rebounds.

We can crudely estimate the threshold necessary for minimizing the peak prevalence. If we know a population’s reproductive number ℛ_0_ and its initially immune fraction *R*^*^ and susceptible fraction *S*^*^ = 1 − *R*^*^, we can determine the peak prevalence.^1^ In the limit of a very long (*D* → ∞) and strong intervention (*c* → 1), at the end of the intervention *S*(*t*^*^ + *D*) ≈ *S*(*t*^*^) and *R*(*t*^*^ + *D*) ≈ *I*(*t*^*^) + *R*(*t*^*^). If *D* is long, but *c* is small enough that we cannot ignore transmissions occurring during the intervention, then we need to correct for the fact that *S*(*t*^*^ + *D*) may be somewhat smaller than *S*(*t*^*^). Accounting for these transmissions further reduces the size of the second peak.

We can use this to estimate when *I*(*t*^*^) will approximate the rebound. As this is not strongly dependent on duration, or *c*, this explains why the optimal threshold for peak prevalence does not vary much in Figures 3(b) and (e).

It is worth highlighting that in related recent work [30] showed that the penalty for making a small error in the *timing* of the intervention is larger if it is too late compared to too early. As we see in our figure, the error as a function of the threshold *I* + *R* appears roughly symmetric, but because the optimal intervention time often occurs while the epidemic growth is increasing, this means that being a little too late means a larger error in *I* + *R* than being a little too early.

#### 3.1.3 Impact on timing of infections

The impact on the timing of an emerging epidemic is an additional factor that plays an important role. If we anticipate rapid development of new treatments or interventions, then this may be more important than reducing the anticipated peak prevalence or total fraction infected.

As we noted for the attack rate and the peak prevalence discussions, for very early interventions (for which *I* + *R* is very small), the entire epidemic curve shifts in time.^2^. However, as the threshold increases and we start to see an impact on the final attack rate *R*(∞) and peak prevalence *I*_max_, we also see an additional impact on the average time of infection. Unlike the other targets, a later intervention tends to have an decreased impact because more of the infections have occurred earlier.

In a real-world context, we anticipate that the model may overstate the delay from a very early intervention if there is significant transmission outside the population of interest. In a setting where the disease is spreading outside the population, the reduction of infections within the population during the intervention may be immediately negated by new transmissions from outside, which are likely to be increasing. So the effect to delay the epidemic is largest if accompanied by reduction in transmissions from outside. However, in a setting where the disease is not well-established outside the population (as occurred in China early in the COVID-19 pandemic), or travel from outside can be restricted, a major effort at early time may significantly delay the eventual epidemic.

### 3.2 Weakly-coupled metapopulation model

We now consider a more realistic population which consists of coupled subcommunities, effectively a metapopulation model. We again consider one-shot interventions that either target the entire population at once (synchronous interventions) or that target individual subcommunities at different times (asynchronous interventions). If they were strongly coupled, the epidemics would be synchronous [4, 12]. So the single well-mixed population results would carry over. Our focus is on weakly-coupled subcommunities.

A typical plot of the prevalence level in each subcommunity is shown in Figure 4 in the absence of intervention. The epidemic starts in subcommunity two but it then spreads to the others. The entries of the cross-infection/mixing matrix are generated following the description in Section 2.2, and the specific mixing parameters are given in the Appendix.

**Figure 4:**
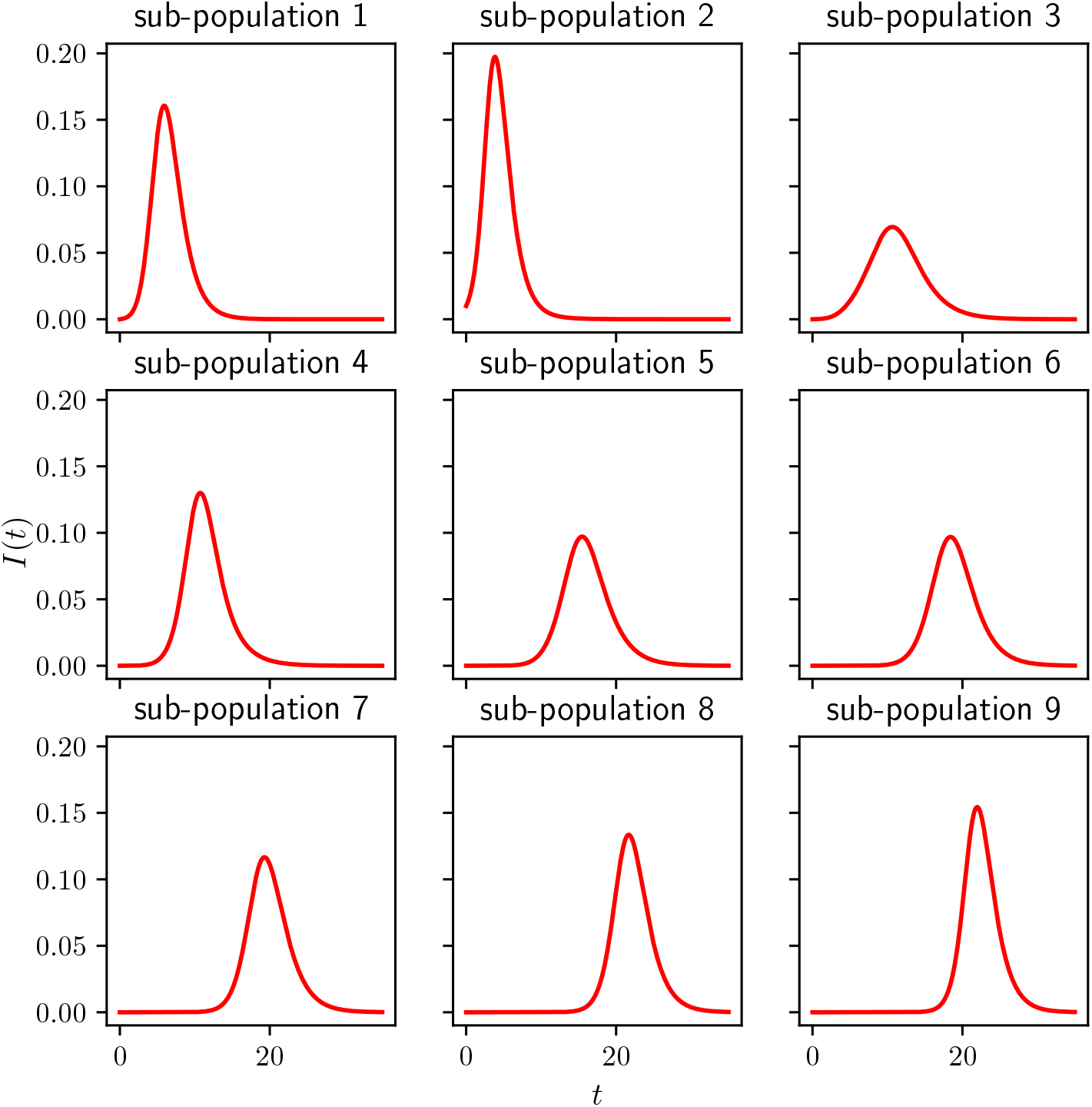
Example of an epidemic spreading across 9 subcommunities with different contact rates (see the Appendix A.2 for the precise mixing matrix *B*). The epidemic starts from subcommunity 2 and it is run for *T* = 35 units of time. *γ* = 1 for all subcommunities. With no control the attack rate or final epidemic size is 0.744.

As before we consider the impact of intervention on attack rate, peak prevalence, and peak timing. The overall effect of interventions is qualitatively similar to that of the single-population model. However, we find that asynchronous interventions that target each sub-community significantly outperform synchronized interventions that begin when either the first subcommunity reaches a threshold or the global infection crosses a threshold.

For synchronized interventions, the overall impact is smaller, and the best outcomes are not driven by the actual threshold value. Rather they result from the intervention being timed to have significant impact on multiple communities, or optimally delaying the spread between communities. Consequently, the ideal times for this will depend on the parameters for between-community transmission, and are likely to be population-dependent.

Because the epidemics subcommunity may not be synchronized across subcommunities, this means that when a synchronous intervention is applied some subcommunities may have already completed their epidemic, or others have not yet begun. Interventions that are based on the first population to reach a threshold may not be valuable if the particular intervention is most effective if it disrupts patterns that do not appear until the first subcommunity has effectively completed its epidemic.

In our results, we consider 100 simulated populations consisting of 9 subcommunities, whose contact structure is generated from the random process described in Section 2.2. For most results we present only the average behavior. We note that this aggregation may hide important behavior from individual simulations [21]. However, except where noted, our averaged results are qualitatively similar to individual populations. Where we look at an individual simulation, we use the specific population of Figure 4.

#### 3.2.1 Impact on attack rate

Our primary observation about the attack rate is that interventions acting at different times for each subcommunity are substantially more effective than synchronized interventions.

The smallest values of the attack rates are achieved when control acts independently in each subcommunity meaning that as soon as *I*_*i*_ + *R*_*i*_ crosses a threshold, the one-shot control is switched on in subcommunity *i*. This is done independently of whether the efficacy or duration of control is kept fixed, while the other is varied, see Figure 5(a,d). Typically, as in the case of a single population, there seems to be a clear optimal threshold value which leads to the smallest attack rate. Applying the control too early or too late leads to higher attack rates. Fixing the threshold value and increasing the duration of control, see Figure 5(a), or the strength of control, Figure 5(d), leads to smaller attack rates. Both strength and duration of control have no significant impact on the attack rate if the intervention is too early or too late.

**Figure 5:**
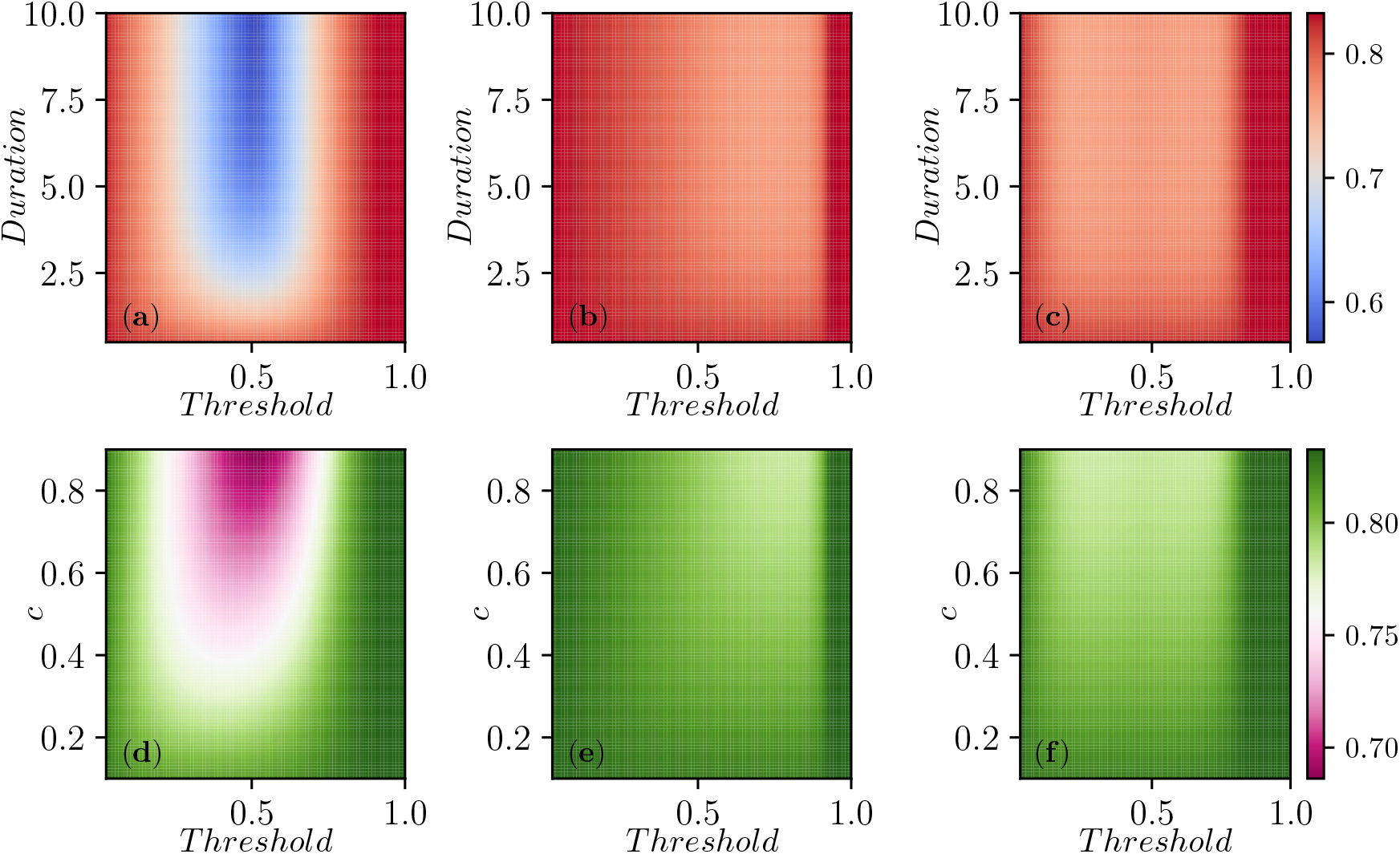
Contour plots showing the average attack rate (final epidemic size) over 100 simulated populations for each set of parameter values. In the first row *c* is fixed and the duration of control varies on the vertical axis, while in the second row duration is fixed and *c* varies. Each column corresponds to one of the three strategies: (a,d) intervention in each subcommunity, (b,e) global intervention when the first subcommunity breaches the threshold, and (c,f) global intervention at global threshold for a population consisting of 9 subcommunities. In each plot, the *x* − *axis* shows the values that the threshold for intervention can take (from a minimum of 0.05 to a maximum of 0.8). In the first row *c* = 0.8 is constant, while the duration of control varies from a minimum of *T* = 1 to a maximum of *T* = 10. On the second row instead, the duration of control is kept fixed at *T* = 2, and the values of *c* varies from *c* = 0.1 to *c* = 0.9. The recovery rate is *γ* = 1 for all subcommunities. In all cases, if the threshold is set too large the intervention is never implemented. The two synchronized interventions can be approximately mapped to one another by noting the largest *I*_*i*_ + *R*_*i*_ at the time the global *I* + *R* reaches a given threshold. The subcommunity threshold gives more resolution at small values while the global threshold gives more resolution at large values.

Figure 6 shows how the best one-shot control works when the optimal threshold for fixed control efficacy and duration is implemented. As expected, this plot confirms that intervention happens close to the peak of the epidemic in each subcommunity so secondary waves of infection are heavily suppressed.

**Figure 6:**
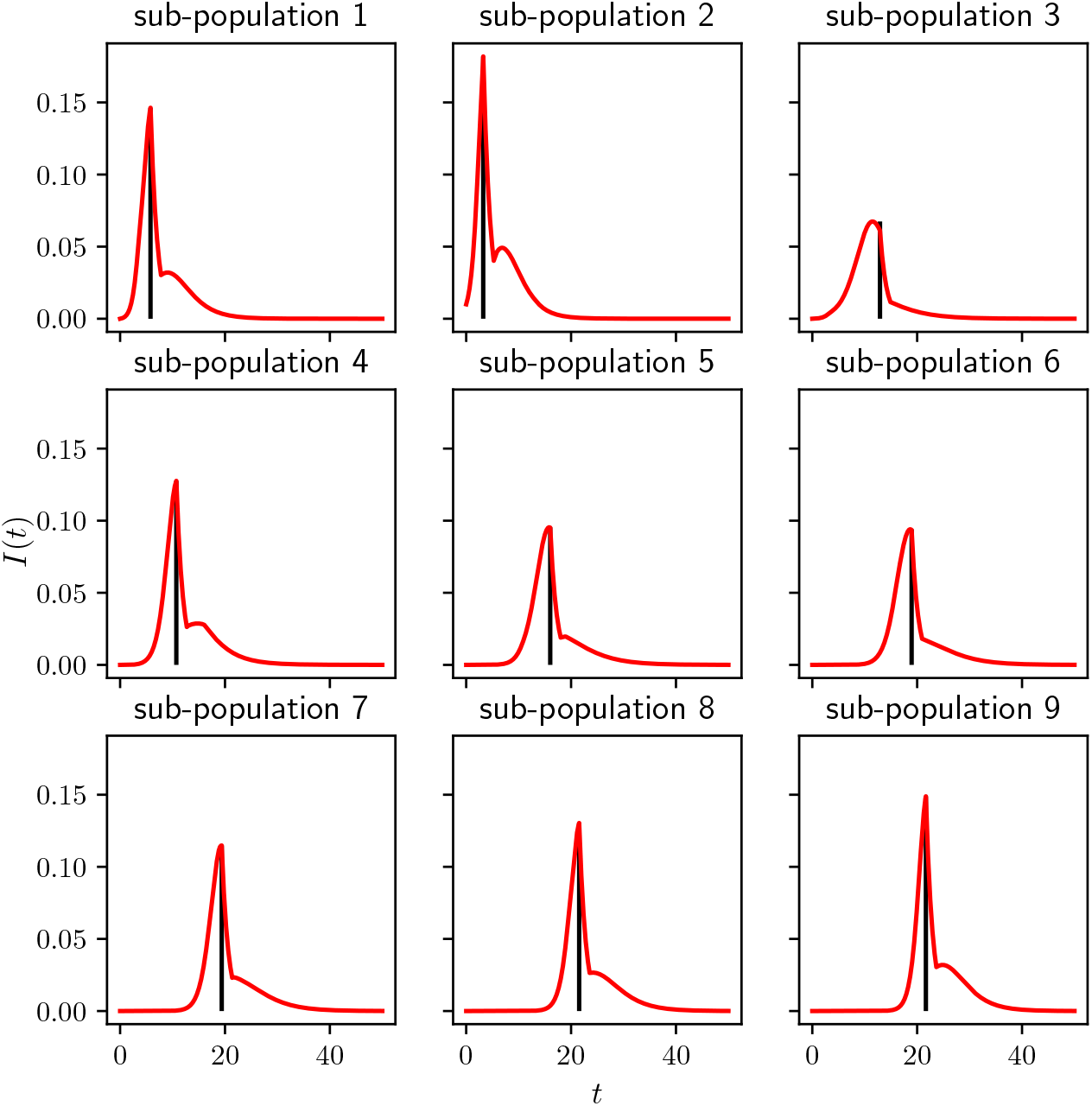
Illustration of best control strategy (i.e. smallest attack rate) (controlling subcommunities individually but using the same threshold for each) when efficacy and duration of control are fixed at *c* = 0.8 and *D* = 2, respectively. It turns out that the optimal threshold is close to (0.4). This combination represents the point (0.4, 2) in Fig 5 panel (a), or equivalently the point (0.4, 0.8) in panel (d). With this strategy, we find that *R*(∞) goes from *R*(∞) = 0.75 to *R*(∞) = 0.63. If we increase control duration from 2 to 10 we would achieve a further reduction to *R*(∞) = 0.44. The vertical black lines show the onset of control.

The impact of the synchronized intervention based on the global level of *I* + *R*, see Figure 5(c,f), or on the first subcommunity to reach a threshold, see Figure 5(b,e), are much smaller than asynchronous interventions. This is because when it is implemented in the synchronous case, some communities have already completed their epidemic while others have not yet begun. So there is less overall impact (see the asynchrony in Figure 4).

When the intervention is based on the first time a subcommunity crosses a threshold, we find that the optimal thresholds are at relatively large values. This suggests that the value of the synchronized intervention comes from disrupting transmission when the disease is spreading in multiple subcommunities.

Under the synchronous intervention scenario, we also see some surprising behavior where there are multiple local maxima for the specific metapopulation used in Figure 4 (not shown in Figure 5). This effect is because the timing aligns with different outbreaks. If we intervene at one time, we may have a big impact on one subcommunity, and if we miss that window, it is best to wait until another subcommunity begins to have an outbreak. This effect disappears in the aggregated data of Figure 5 because the specific ideal timing is a consequence of the randomly chosen parameters of each population.

#### 3.2.2 Impact on peak prevalence

Here we look atthe effect of the intervention on the peak prevalence, that is the maximum value of 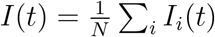 during the time course of the epidemics. As with the attack rate, our primary observation for the peak prevalence is that it is significantly reduced by targeting based on the individual subcommunity.

Figure 7 shows the average of the peak prevalence across the same 100 populations as Figure 5. Perhaps not surprisingly, Figure 7 is qualitatively similar to Figure 5. The most impact is through having interventions occurring when the individual populations reach a threshold. The optimal choices for intervention come earlier in the epidemic. We still observe that if the intervention is too soon or too late then there is no significant reduction in peak prevalence.

**Figure 7:**
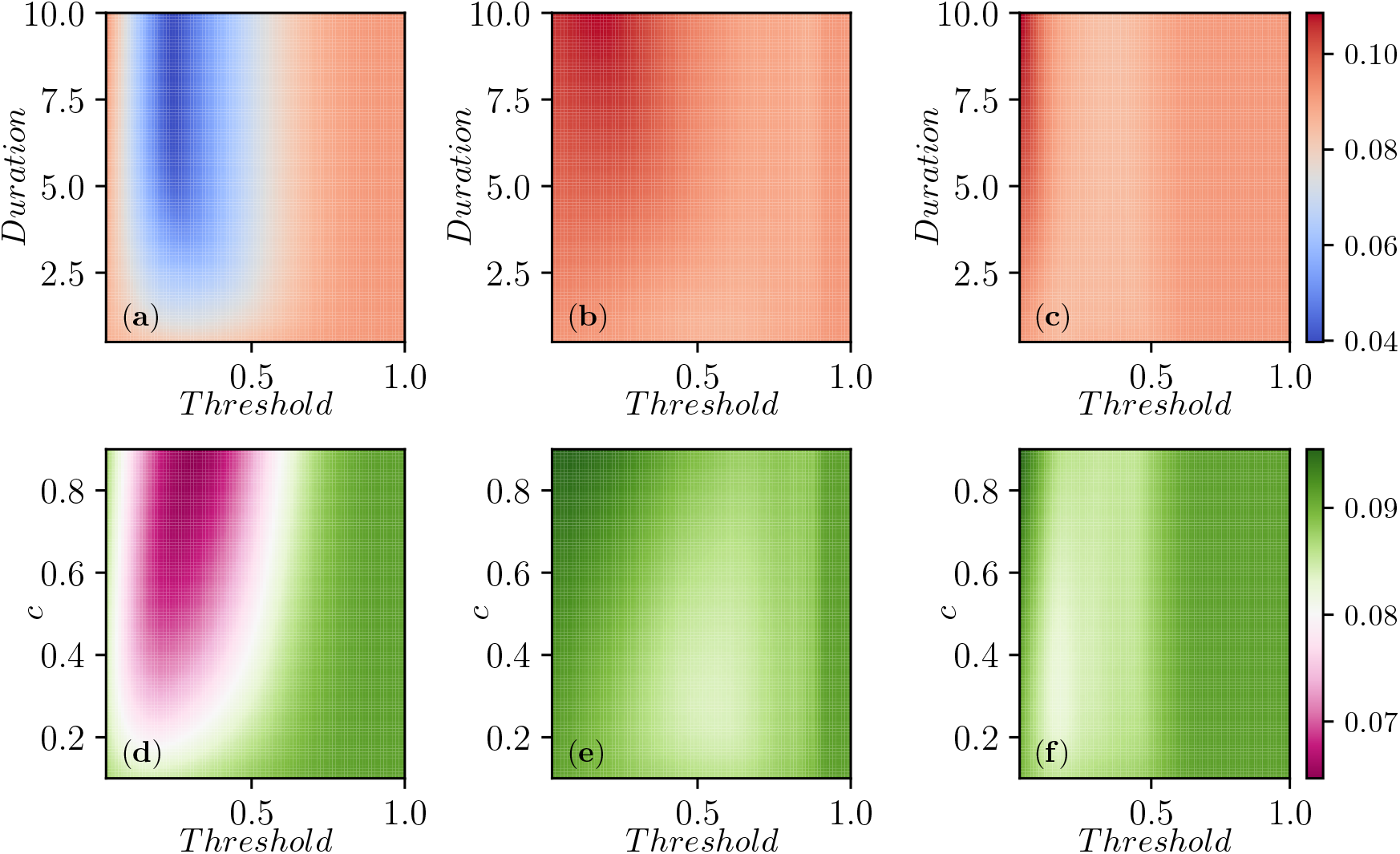
Contour plots of the peak prevalence 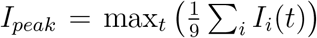, averaged across 100 simulated populations each with 9 subcommunities. Control strategies and setup the same as in Figure 5.

In Figure 7 (a,d) the optimal threshold for intervention is relatively early, this is in line with the trend observed in Figure 2 for the single population case. We should wait until some immunity builds up before intervening, so that the rebound in each population is muted.

For our two synchronized strategies, the effectiveness is much less, because the overall peak prevalence is related to how the individual subcommunities’ peaks align, and different details of intervention timing, combined with the random parameters of the simulation, can make the individual peaks align or not.

Interestingly, if we look at an individual simulations, there are thresholds which yield significantly larger improvements in the peak prevalence than we see in the aggregated data. This is because in each simulated population, the relative timing of the epidemics subpopulations depends on the system parameters. In a weakly-coupled metapopulation model with relatively few subcommunities, the global peak prevalence is likely to occur when multiple subcommunities happen to be aligned. This is highly sensitive to parameters, and so the optimal intervention time will vary.

#### 3.2.3 Impact on epidemic timing

When we investigate the average time of infection, we see that targeting the intervention at each subcommunity is again the most effective. In general the interventions need to be implemented very early in the epidemic.

Most of the impact comes from slowing the epidemic in the initial subcommunity. A delay in the initial place of introduction results in a delay in all subcommunities. Once the intervention is no longer in place in the initial subcommunity it begins to grow and spill over into other communities. If other subcommunities wait until then to begin their response, they gain some benefit. However, once they stop, they face rapid reseeding from the initial subcommunity. So the main benefit comes from the initial subcommunity’s actions. When we use a synchronized intervention, the effect is somewhat smaller, but it is not significantly smaller.

In fact, most of the benefit comes from the initial subcommunity engaging in preventative measures. There is relatively little impact on the average time of infection to be gained from the other subcommunities acting early, unless they can maintain a very small spillover rate for a long period through extensive travel restrictions or similar interventions. This suggests that significant benefit may come from a hybrid strategy which focuses on delaying infections out of the initial subcommunity while other locations focus their interventions on optimizing peak prevalence or attack rate.

## 4 Discussion

We have considered the impact of a single one-shot limited duration impact on the spread of an infectious SIR disease, in both a single well-mixed community and in a weakly-coupled metapopulation model.

We have found that in a single well-mixed population, an intervention at the first hint of infection is best for delaying infections. An intervention that waits until the epidemic is well-established but still well short of the peak is best to reduce the peak prevalence of the epidemic. An intervention (whose duration would be *D*) that starts a little less than *D* units of time before the peak would otherwise be reached is best to reduce the total number of infections.

In a weakly-coupled metapopulation model, we find qualitatively similar results. The best strategy to reduce either the peak prevalence or the total number of infections, a strategy that times the interventions asynchronously to when each subcommunity reaches a threshold rather than being synchronized to when the global average crosses a threshold or when the first subcommunity crosses a threshold.

For delaying the average time of infection, the most important detail is that the sub-community with the first introduction responds as quickly as possible. Whether the other sub-communities respond immediately or delay their response until more infections are present within the subcommunities has a smaller effect.

### 4.1 Limitations

Our results are somewhat limited by the assumptions we have made to produce a tractable problem.

We assume that behavior responds immediately to changes in interventions. In reality, behavior may change prior to an intervention being implemented. Additionally adherence may drop as the intervention continues, and some adherence to the intervention may remain even once the intervention is removed.

The assumption that the individuals are largely homogeneous may lead to pessimistic predictions of when the herd immunity threshold is reached. In the presence of significant population heterogeneity, there is evidence suggesting that the herd immunity threshold would be reached earlier, and the epidemic could proceed significantly faster [16, 7]. Our qualitative predictions remain robust, but the timings would need to move sooner.

We must think critically about what constitutes a one-shot intervention. Whether an intervention can be maintained may depend on context. Early estimates of case fatality rate (not to be confused with infection fatality rate) of COVID-19 ranged from 0.7% in China outside of Hubei province to around 2% in much of the world, to around 5.8% in Wuhan [35]. These estimates were affected by the proportion of cases identified (leading to uncertainty in the denominator), and whether the health system was over capacity (which would increase the death rate leading to uncertainty in the numerator). True infection fatality rates appear to lie between 0.5% and 1%, with many estimates closer to 1% than 0.5% [28, 34, 31]. With such high fatality rates, our tolerance for drastic interventions should increase. Thus an intervention that would be considered one-shot for the 2009 H1N1 pandemic which had a significantly lower fatality rate [36] might be considered sustainable for the COVID-19 pandemic.

In deciding whether an intervention is sustainable policy makers could formulate an answer to this question: “Assume you impose the intervention now, and infection rates remain the same or higher in the future, and would increase if they were dropped, would you be willing to maintain the intervention in place?” If so, then the intervention is sustainable. If the answer is “no”, and the intervention will be abandoned at some future time regardless of the new infection profile, then this is a one-shot intervention, and it should be held in reserve until it will have maximal impact.

We have ignored logistical challenges that might be associated with implementing the intervention separately for each subcommunity. On a large scale (e.g., states within a country or cities within a state) we anticipate that this is logistically feasible, while on a small scale (e.g., suburbs in a city) it is more likely that the epidemics will be synchronized and this benefit is small compared to the logistical challenges.

### 4.2 Policy Implications

Our observations have a number of important policy implications for an epidemic which is sufficiently established that elimination is not a goal. Primarily:

- To reduce the attack rate of an epidemic, a one-shot intervention should be introduced shortly before the epidemic peak.
- To reduce the peak size of an epidemic, the intervention should be late enough to allow significant immunity to develop, but early enough to allow a substantial rebound after the intervention.
- To delay infections as much as possible, the intervention should be implemented early on.
- In a population made up of many weakly-coupled subcommunities, interventions should be asynchronous.

Because they require different timings, the three goals we have considered are in conflict. The benefits of reducing the total number of infections are clear, and if health care capacity is threatened, the benefits of minimizing the peak prevalence are also clear. It is less obvious that delaying infections may help. However when there is an expectation that improved treatments and improved inteventions may be developed, a delay is likely to be the best available option.

An additional benefit of delaying the epidemic is observed when we have an intervention, such as testing & tracing, which does not scale well. As the number of infections grows ebyond the testing capacity, the effectiveness per infection goes down. At low infection levels these interventions may be enough to suppress transmission. However, if levels get too high, then a quick intervention that causes infections to drop may be more effective than an intervention that waits until the optimal time to minimize the size.

Although we have focused on three distinct goals which lead to different optimal strategies, the ideal goal is likely to be a combination of these effects. So the timing will need to respond to different pressures. If, for example, the goal is to keep the peak prevalence below a certain value while minimizing the maximum prevalence, then the ideal strategy would let as much immunity develop in the population as possible before the prevalence limit is reached and then intervene (this is of course only feasible if there is a time that can keep prevalence below that level). Delaying the intervention until this point would mean that the second peak would be lower. If the goal is instead to keep the peak prevalence below some level while maximizing the delay of infections, then the intervention would be sooner and timed so that the second peak would reach the target prevalence.

We finally note that in a population made up of weakly-coupled subcommunities whose epidemics will not be synchronized, the ideal intervention might be to react strongly and immediately in the first subcommunity where the infection begins to spread. This can provide protection to the other communities and significantly delay the spread. Once it spreads beyond that initial subcommunity then the focus may turn to minimizing the peak prevalence or the attack rate.

## A Mathematical Analysis

In this section we provide mathematical analyses of the single population model to support our results for reducing attack rate and peak prevalence.

### A.1 A Phase-plane based analysis

Because *S* + *I* + *R* = 1, we can fully specify the current state and the future dynamics by knowing *S* and *R*, in which case *I* = 1 − *S* − *R*. It will be useful to use this to explore the dynamics of an epidemic and the impact of an intervention.

In Figure 10 we show how *S*(*t*) and *R*(*t*) evolve together in time for three values of *R*(0) (0.5, 2, and 4) and for several different initial conditions. For a given point (*S*_0_, *R*_0_), the value of *I* at that time is given by the horizontal (or vertical) distance to the diagonal line *S* + *R* = 1.

**Figure 8:**
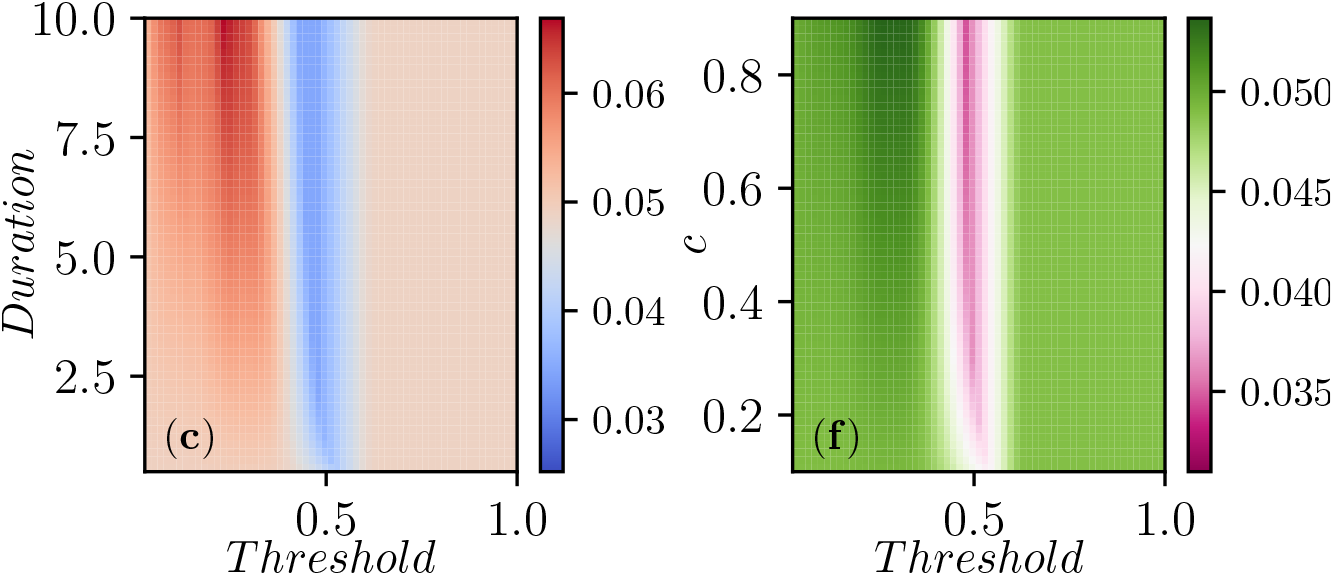
Contour plots of the peak prevalence, *I*_*peak*_, that is the maximum value achieved by 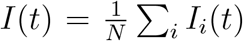 during the time-course of the epidemic. Control strategies and setup the same as in Figure 5.

**Figure 9:**
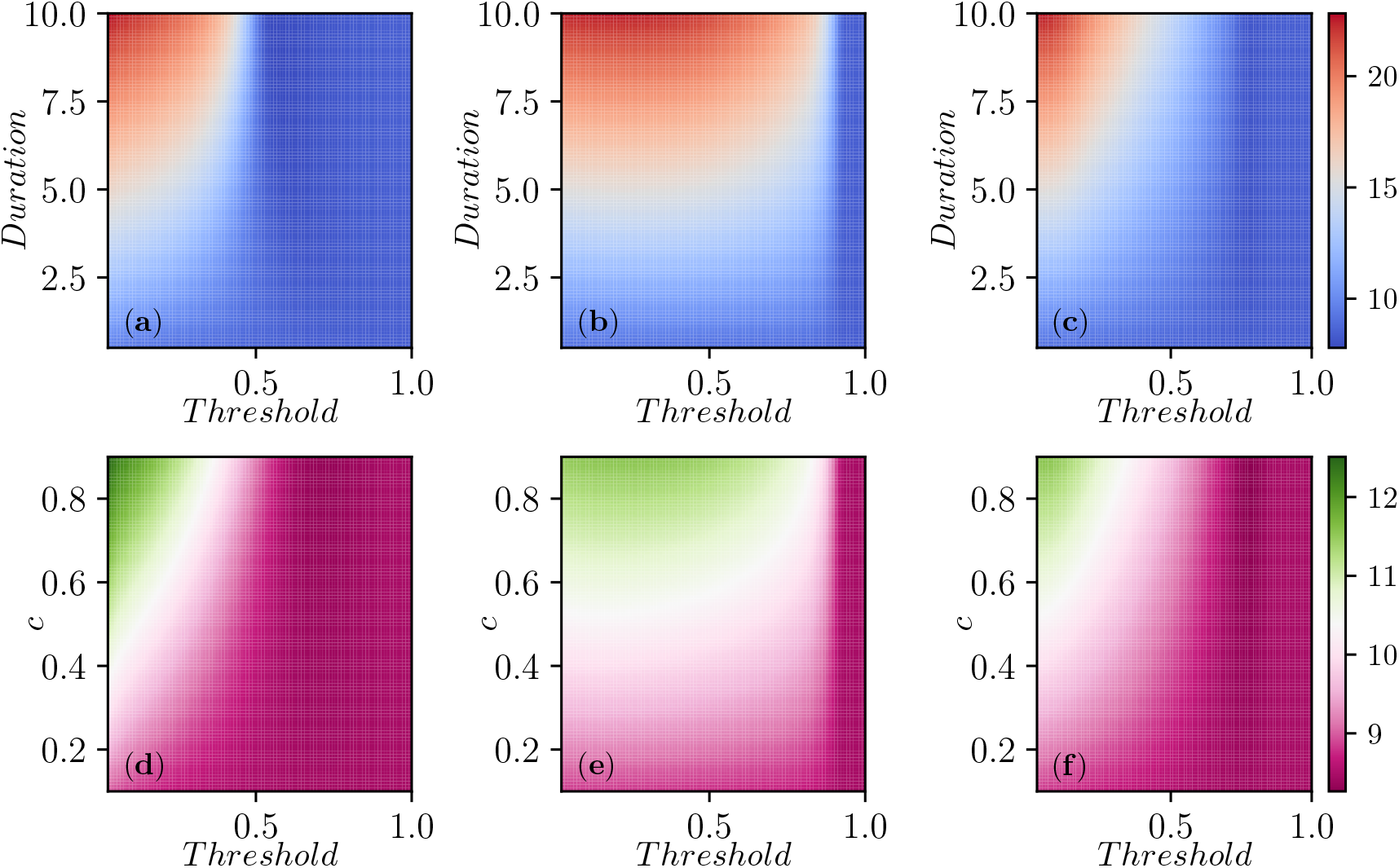
Countour plots of the global mean infection time, defined as 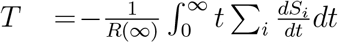, averaged over 100 simulations. In terms of control strategies and parameter values the same setup as in figures 7 and 5 are used.

**Figure 10:**
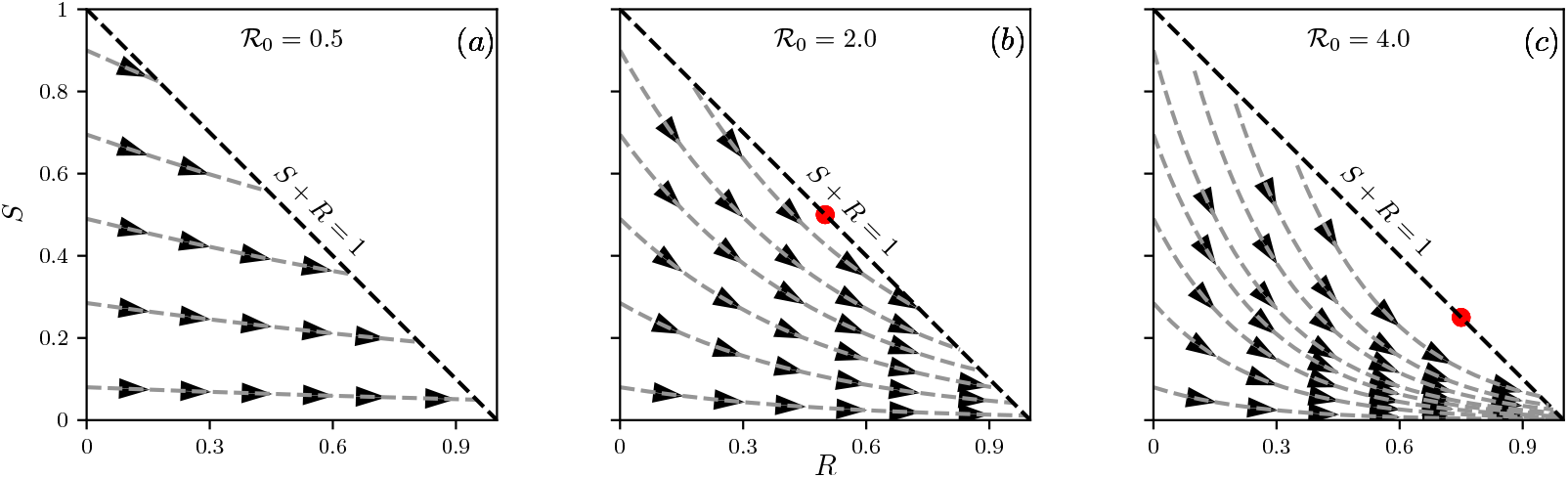
We plot *S*(*t*) versus *R*(*t*) for ℛ_0_ = 0.5 (a), 2 (b), and 4 (c). For given *S*(*t*) and *R*(*t*), the proportion infected is *I*(*t*) = 1 − *S*(*t*) − *R*(*t*), which equals the vertical or horizontal distance from the point (*R*(*t*), *S*(*t*)) to the line *S* + *R* = 1. The curves and arrows show how a solution to System (1) evolves in time. At points *S* > 1*/*ℛ_0_ (which occurs only for ℛ_0_ > 1) orbits move farther from the diagonal, representing an increase in *I*. Note that the velocity an orbit is traversed varies depending on location, and goes to zero close to *S* + *R* = 1. Red dots in panel (b)-(c) indicate the point 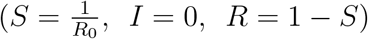.

In the figure, we can see that if *S* > 1*/*ℛ_0_ (which is only possible if ℛ_0_ > 1), then the horizontal distance from the orbit to the *S* + *R* = 1 line is increasing as the orbit moves forward. In other words, *I* is increasing. Once *S* < 1*/*ℛ_0_, the distance decreases and eventually goes to 0.

Using these orbits, we can investigate the impact of an intervention, as shown in Figure 11. We follow *S* and *R* along an orbit. When we turn on the one-shot intervention at time *t*^*^, it no longer follows the original curve. Instead the change in *S* and *R* follows the paths we would find for (1 − *c*) ℛ_0_, starting from the point (*R*(*t*^*^), *S*(*t*^*^)). It follows this curve until reaching (*R*(*t*^*^ + *D*), *S*(*t*^*^ + *D*)) when the intervention is halted. It then follows the curves for the original ℛ_0_, but starting from this new point.

**Figure 11:**
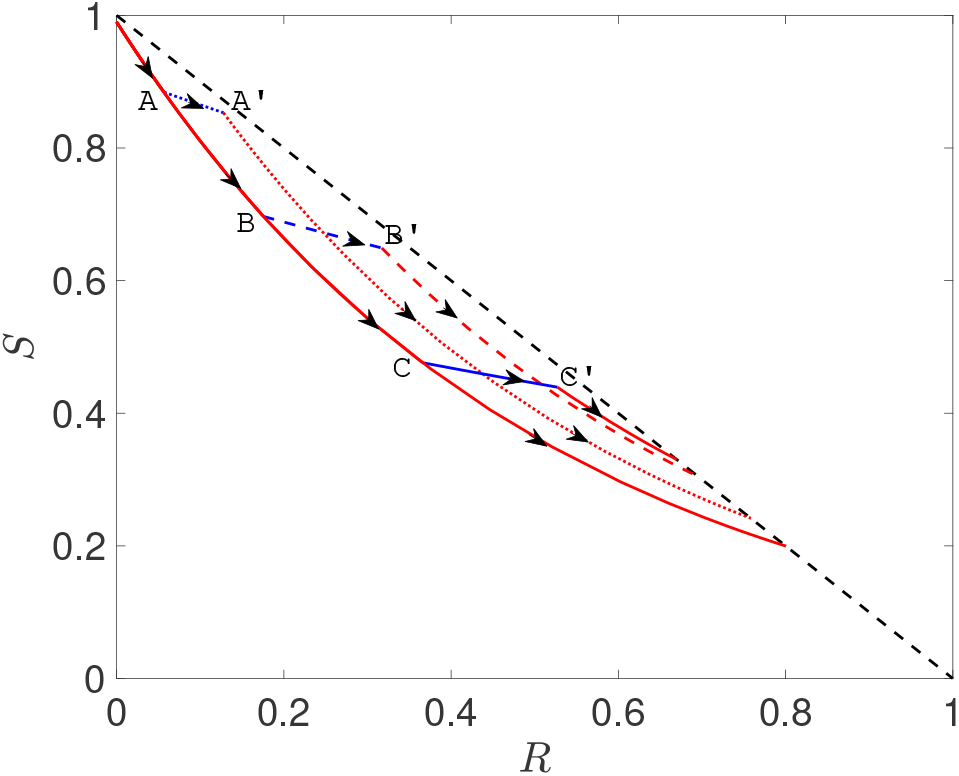
(S,R) phase portrait (arrows indicate growing time) based on an SIR model in a single population with *β* = 2, *γ* = 1 (giving ℛ_0_ = 2) and initial condition *I*(0) = 0.01. The plot shows a trajectory with no control (continuous red line) as well as three other trajectories where *β* = 0.5 for a time period of length *D* = 2 but with the intervention setting in only once *I* + *R* goes past 0.1 (partially dotted line), 0.3 (partially dashed line) and 0.5 (continuous broken line), respectively. Control for the three different scenarios sets in at the points denoted by A, B and C and control ends at A’, B’ and C’, respectively.

Note that there is a point (*R, S*) = (1 − 1*/* ℛ_0_, 1*/* ℛ_0_) at which separates the points on the line *R* + *S* = 1 from which an epidemic could start from the points at which epidemics finish. The closer an orbit is to this point, the smaller the attack rate.

So for ℛ_0_ = 2, a temporary intervention gives us a way to move from one of these curves in the ℛ_0_ = 2 plot to another. We see this in Figure 11. The timing of the intervention determines which of the orbits the system lands on.

In this context the goal of reducing the attack rate is equivalent to ensuring that the intervention shifts the curve to an orbit as close as possible to (*R, S*) = (1−1*/* ℛ_0_, *S* = 1*/* ℛ_0_). Reducing the peak prevalence is equivalent to ensuring that the orbit remains as close as possible to the line *S* + *R* = 1.

#### A.1.1 Attack rate

If our goal is to minimize the number of infections, we accomplish this by having the curve (*R*(*t*), *S*(*t*)) land on an orbit that is as close to (*R, S*) = (1 − 1*/* ℛ_0_, 1*/* ℛ_0_) as possible given the constraints on the intervention.

Typically we have to wait until the curve has moved closer to the desirable orbits before implementing the intervention. Implementing the intervention early, see the dotted line in Figure 11 means that at the end of the intervention there is still a large pool of susceptibles which are at risk of becoming infected. Crossing from A to A’ simply puts the epidemic on a slightly different trajectory but the attack rate is very close to the case with not control.

An intervention at a later stage, see dashed line, improves the final outcome resulting in an attack rate that is smaller when compared to the case of no control. Finally, the continuous broken line shows an almost optimal intervention with a further small reduction in the attack rate.

In general, the intervention that will get us closest to the optimal value occurs when the original curve is close to, but has not yet reached, the largest value of *I*, which occurs when *S* = 1*/* ℛ_0_. As the effectiveness of the intervention increases, the orbits it follows during the intervention become more horizontal. For very effective interventions, this suggests we should wait until very close to the epidemic peak, while for less effective interventions (which will slope downwards more), we will want to implement them somewhat sooner.

#### A.1.2 Peak prevalence

For peak prevalence, the goal is to keep the curve as close as possible to *S* + *R* = 1. The longer we wait to implement the intervention, the closer the final orbit is to *S* + *R* = 1, but the farther the original orbit moves from the line. With this in mind it becomes clear that the optimal *t*^*^ to reduce peak prevalence is smaller than the optimal value to reduce attack rate.

### A.2 The mixing matrix

The cross-infection between subcommunities is modelled by *B* = (*β*_*ij*_)_*i,j*=1,2,…*N*_, where *β*_*ij*_ represents the rate at which infectious contacts are made from subcommunity *i* towards susceptible individuals in subcommunity *j*.

We implement a weak coupling by joining the population in a linear fashion: population *i* is only connected to population (*i* − 1) and (*i* + 1). The first and the last populations only connect to the second and the pen-ultimate population, respectively. The entries for the coupling/mixing matrix are generated as follows. On the main diagonal, the *β*_*ii*_ values are set to 2 + (Unif(0, 1) − 0.5). Off-diagonal entries are set to Unif(0, 1 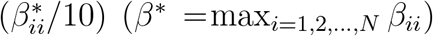 and represent a scaled and randomised version of the largest entry on the main diagonal. This yields an R_0_ above 2, comparable to current estimates for COVID-19. For most of our results we aggregate over 100 distinct simulated populations. However, in figures 4, 6, and 8 we use a single realization of the metapopulation model. For this we take the mixing matrix

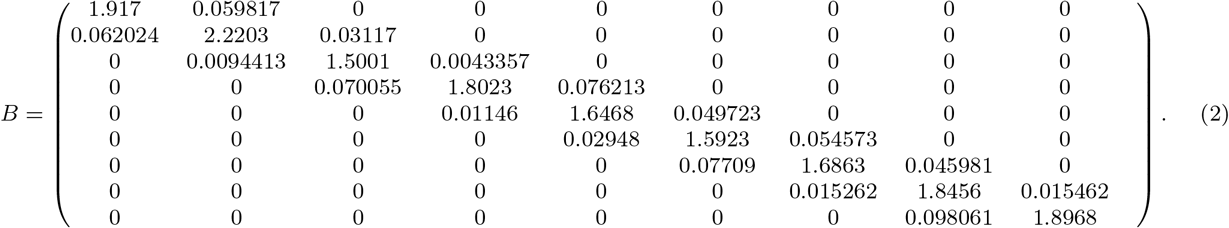

## Data Availability

The model is entirely simulation based. No data has been used.

## Footnotes

1 There is an analytic formula for peak prevalence 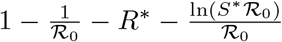 but for our purposes we just need to recognize that ℛ_0_, *R*^*^ and *S*^*^ are sufficient to determine it.

2 Of course if the disease is eliminated locally which is more likely with a small threshold, then the next peak depends on frequency of reintroduction which we do not consider

## Notes

### Competing Interest Statement

The authors have declared no competing interest.

### Funding Statement

This work was supported by the authors' institutions and by The Leverhulme Trust, RPG-2017-370

### Summary of Updates

Revised and updated, with more discussion about the impact of delaying epidemics.

